# Enhanced production of eicosanoids in plasma and activation of DNA damage pathways in PBMCs are correlated with the severity of ancestral COVID-19 infection

**DOI:** 10.1101/2023.09.14.23295549

**Authors:** Jeffrey A. Tomalka, Anna Owings, Michelle Galeas-Pena, Carly G.K. Ziegler, Tanya O. Robinson, Thomas G. Wichman, Hannah Laird, Haley B. Williams, Neha S. Dhaliwal, Steven Everman, Yousaf Zafar, Alex K. Shalek, Bruce H. Horwitz, Jose Ordovas-Montanes, Sarah C. Glover, Yann Gibert

## Abstract

**Background:** Many questions remain unanswered regarding the implication of lipid metabolites in severe SARS-CoV-2 infections. By re-analyzed sequencing data from the nasopharynx of a previously published cohort, we found that *alox* genes, involved in eicosanoid synthesis, were up-regulated in high WHO score patients, especially in goblet cells. Herein, we aimed to further understand the roles played by eicosanoids during severe SARS-CoV-2 infection.

**Methods and findings:** We performed a total fatty acid panel on plasma and bulk RNA-seq analysis on peripheral blood mononuclear cells (PBMCs) collected from 10 infected and 10 uninfected patients. Univariate comparison of lipid metabolites revealed that lipid metabolites were increased in SARS-CoV-2 patients including the lipid mediators Arachidonic Acid (AA) and Eicosapentaenoic Acid (EPA). AA, EPA and the fatty acids Docosahexaenoic acid (DHA) and Docosapentaenoic acid (DPA), were positively correlated to WHO disease severity score. Transcriptomic analysis demonstrated that COVID-19 patients can be segregated based on WHO scores. Ontology, KEGG and Reactome analysis identified pathways enriched for genes related to innate immunity, interactions between lymphoid and nonlymphoid cells, interleukin signaling and, cell cycling pathways.

**Conclusions:** Our study offers an association between nasopharynx mucosa eicosanoid genes expression, specific serum inflammatory lipids and, subsequent DNA damage pathways activation in PBMCs to severity of COVID-19 infection.

## Introduction

The COVID-19 pandemic has led to a global health crisis, with millions of people infected and hundreds of thousands of deaths worldwide (1). COVID-19 disease is caused by the SARS-CoV-2 virus, which primarily affects the respiratory system, leading to severe respiratory distress and acute respiratory distress syndrome (ARDS) in the most severe cases (2). However, it is now well established that COVID-19 is not just a respiratory disease, but a systemic disease that affects multiple organs and systems in the body (3).

One of the key features of severe COVID-19 infection is an exaggerated inflammatory response, which can lead to a cytokine storm and multi-organ failure (4–7) . Eicosanoids are a group of bioactive lipids that are synthesized from arachidonic acid and play a crucial role in the regulation of inflammation and immune responses (8). Prostaglandins and leukotrienes are two major classes of eicosanoids that are involved in the initiation and propagation of inflammation (9). Recent studies have suggested that dysregulation of the arachidonic acid cascade and increased synthesis of pro-inflammatory eicosanoids may contribute to the pathogenesis of severe COVID-19 infection (10,11). On the other hand, specialized pro-resolving mediators (SPMs) are a group of lipid mediators that are synthesized from omega-3 fatty acids and play a crucial role in the resolution of inflammation and tissue repair (12). SPMs have been shown to have potent anti-inflammatory and pro-resolving effects and may have therapeutic potential in the treatment of COVID-19 (13). SPM concentrations were correlated with both circulating phagocyte activation status and function (14). The inflammatory response in COVID-19 is dynamic and can fluctuate extensively from one day to another (15). Previous research has noted cellular immune dysregulation in COVID-19 (16). Furthermore, the presence of inborn error of immunity may increase the risk of infection and severity of COVID-19 (17).

The impact of severe SARS-COV-2 infection on lipid metabolism is currently not fully understood. However, the number of cardiovascular events and the development of autoinflammatory syndromes which have increased in previously infected individuals suggests that SARS-COV-2 may lead to disordered lipid metabolism (18–20).

Measurement of eicosanoids including SPMs in biological fluids such as nasal swabs, urine, and blood samples has become an important tool for further understanding the role of lipid mediators in disease pathogenesis and development of new therapeutic strategies (21). Recent studies have not only identified temporal clusters of endogenously produced eicosanoids and SPMs in blood suggesting they play a role in host defense and coagulation (22) but has also used measurements of eicosanoids and SPMs to investigate their role in various diseases, cancer, asthma, etc. (23).

Of interest in the study of lipid metabolism and their role in the immune responses are lipoxygenase genes, such as alox5 and alox12/15. Lipoxygenases catalyze the oxidation of polyunsaturated fatty acids (PUFA) and the rate-limiting step in the production of leukotrienes, proinflammatory lipid mediators and SPMs. Herein, we re-analyzed our published ancestral nasopharyngeal COVID-19 sc-RNA-seq dataset (24) for presence of alox gene signatures and based on our finding of increased expression, performed lipidomics on plasma, and bulk RNA sequencing on peripheral blood mononuclear cells (PBMCs) from those ancestral SARS-CoV-2 patient samples. We hypothesized that the levels of these mediators would be higher in participants with more severe infection and also correlate to the severity of the infection.

## Methods

### Study participants

Cohort description has been previously published by Ziegler et all (24). In summary, eligible participants were recruited from to the University of Mississippi Medical Center (UMMC) outpatient clinics, medical surgical units, Intensive Care Units (ICU), or endoscopy units between April 2020 and September 2020 (Supplemental Table 1). The UMMC Institutional Review Board approved the study under IRB#2020-0065. All participants, or their legally authorized representative provided written informed consent. Participants were eligible for inclusion in the COVID-19 cohort if they were at least 18 years old, had a positive nasopharyngeal swab for SARS-CoV-2 by PCR, had COVID-19 related symptoms including fever, chills, cough, shortness of breath, and sore throat, and weighed more than 110 lb. Participants were eligible for the Control cohort if they were at least 18 years old, had a current negative SARS-CoV-2 test (PCR or rapid antigen test), and weighed more than 110 lb. Exclusion criteria for both cohorts included a history of blood transfusion within 4 weeks and subjects who could not be assigned a definitive COVID-19 diagnosis from nucleic acid testing. 10 individuals with COVID-19 were included, both male (n = 5) and female (n = 5). For the Control cohort, 10 participants were included 4 identified as male, 6 as female. The median age of COVID-19 participants was 59.5 years old; the median age of Control participants was 62 years old. Among hospitalized participants, samples were collected between Day 1 to Day 3 of hospitalization. COVID-19 participants were classified according to the 8-level ordinal scale proposed by the WHO representing their peak clinical severity and level of respiratory support required.

### Plasma and peripheral blood mononuclear cells (PBMCs) collection and isolation

Blood samples were collected by trained healthcare provider using 10 mL heparinized tubes. Collectors would don personal protective equipment (PPE), including a gown, non-sterile gloves, a protective N95 mask, a bouffant, and a face shield for the COVID positive samples. For plasma isolation the blood was centrifuged at 1200 xg for 10 minutes at room temperature, then collected and stored in -20°C until analysis. For PBMC isolation Ficoll-Paque PLUS density gradient media was added to SepMate 50 mL tubes and then the blood. This was then centrifuged at 1200 xg for 10 minutes at room temperature. The top layer was transferred to a new 50 mL SepMate tube and then centrifuged at 300 xg for 8 minutes at room temperature. The supernatant was removed and the cells washed with 10 mL of 1x PBS. This was then centrifuged at 300 xg for 5 minutes at room temperature. The supernatant was removed at the cells resuspended in 1 mL of CryoStor CS10 freezing media. The PBMCs were then slowly frozen using Mr. Frosty in -80°C and then transferred to liquid nitrogen until analysis.

### Lipid composition measurements by Mass Spectrometry

Lipid extraction was done on each plasma sample. Samples were homogenized in 500 µL of 10% methanol in water and 200 µL of the homogenates were extracted by a modified BUME method. Extracts were dried and saponified using a 1:1 MeOH:KOH solution at 37°C for 30 minutes. Fatty acids were extracted by a bi-phasic solution of acidified methanol and isooctane, derivatized using PFBB, and analyzed using SIM by GC-MS on an Agilent 6890N gas chromatograph equipped with an Agilent 7683 autosampler. Fatty acids were separated using a 15m ZB-1 column (Phenomenex, Le Pecq, France) and monitored using SIM identification (25,26).

### Lipidomics analysis

To analyze lipidomic profiles, we first performed Principal Component Analysis (PCA) to visualize variance in our data and whether this variance was associated with difference in clinical outcome. PCA uses dimensional reduction to identify the largest drivers of variance across the entire dataset, termed principal components (PC), and then generates a per sample score, reflective of expression of the components, for each PC. PC1 and PC2, the two largest drivers of variance, are then mapped onto a 2-D plot. This allows the researcher to investigate high dimensional datasets in the two dimensions which most accurately account for the variance in the data. For heatmaps, raw metabolomic data was log2 transformed. A value of 1 was added to all raw data so that undetected (0) samples could still be log2 transformed. The package ‘pheatmap’ was then used to generate row normalized heatmaps based on the log2 transformed lipidomics data. Samples were clustered using Euclidean distance and annotated based on key clinical parameters. Significantly differentially expressed metabolites were assessed using Mann-Whitney U-tests. An asterisk (*) was used on heatmaps to identify lipid metabolites which were statistically significantly different. Spearman correlations were run to identify lipid metabolites which were positively or negatively correlated with WHO disease severity score. Network visual was made in Cytoscape by importing lipid metabolites, rho of each metabolite compared to WHO disease severity, and the corresponding p-value for each correlation (27). A network was then made around WHO disease severity in which the color (red) of the connecting line reflects the degree of correlation and the thickness of the connecting line represents the -log10 transformation of the p-value. The thicker and darker red a connecting line, the stronger and more significant the correlation.

### RNA isolation and bulk RNA Sequencing

PBMCs from both control patients (n=10) and COVID-19 infected patients (n=10) were used. After isolating the PBMCs the RNA was isolated using RNeasy Mini Kit (Qiagen) according to the manufacturer’s instructions. RNA sequencing was performed by the UMMC Molecular and Genomics Core Facility (www.umc.edu/genomicscore). Libraries were developed using the Illumina TruSeq mRNA Stranded Library Prep Kit (Set-A-indexes), quantified with a Qubit fluorimeter (Invitrogen), and assessed for quality and size using the QIAxcel Advanced System. Samples were pooled into a single library and sequenced using the NextSeq 1000/2000 P2 Reagents (200 cycles, paired-end 100bp) on the Illumina NextSeq 2000 platform. Quality of the sequenced reads was assessed using the Illumina BaseSpace Cloud Computing Platform. FASTQ sequences were aligned to the Human genome [GRCh38 (884148)] using an RNA-Seq Alignment Application (DRAGEN v3.7.5). Differential expression was determined using the DRAGEN Differential Expression Application (v4.0.3) and DESeq2. Gene expression differences are expressed as log2 fold change (ratio) and q > 0.05.

### RNA-seq analysis

Sequenced reads were assessed for quality using FastQC. We identified multiple samples which showed potential QC issues based on GC content, duplication levels and overrepresented sequences. None of these were deemed likely to impact the efficiency of sequencing alignment so all samples were processed further. Raw FASTQ sequence files were used to align reads to the GRCh38 human genome using the Spliced Transcripts Alignment to a Reference (STAR) method (28). All samples had >75% of reads mapped to unique locations in the human genome. STAR was also used to generate the raw read counts. One sample, COVID-19+ sample #394, was removed from further analysis following STAR alignment as it was overrepresented with genes with low read count values suggesting some error in isolation, processing or sequencing. Raw read counts were then normalized using the trimmed mean of M-values (TMM) method and QC’d again using FastQC (29). All samples passed QC after normalization of read counts. Differential gene expression was then performed on normalized read counts using the voom method (30), as deployed in the package limma (31). Significantly differentially expressed genes (DEGs) were identified for the comparisons between 1) COVID-19 high disease severity and COVID-19 low disease severity groups and 2) COVID-19 samples vs. healthy control samples. All DEGs pass an adjusted p-value of <0.05. A combination of PCA plots, volcano plots and row normalized heatmaps were used to visualize DEGs between groups of interest. Networks of DEGs were created to show their association to one another using GeneMANIA. Pathway enrichment analysis to identify immunological and cellular pathways enriched for DEGs was performed in ClueGO. Spearman correlations were used to identify the transcriptional genes and pathways (ClueGO) which positively and negatively correlated with: 1) the magnitude of disease severity by WHO score and 2) the levels of circulating Eicosapentaenoic Acid (EPA), a key inflammatory lipid identified in our analysis.

### GeneMANIA to visualize differentially expressed genes

For granular identification and listing of differentially expressed genes, the GeneMANIA application in Cytoscape was used to make networks (32). For each set of differentially expressed genes of interest, the list was imported into GeneMANIA. A network was then built based off of the list of genes, connecting them based on published/known Co-expression, Co-localization, Genetic Interactions, Pathway, Physical interactions, Predicted, and Shared protein domains. This visual network identifies all the genes by name and shows how they are known to be connected to one another.

### Performing network-based pathway enrichment using ClueGO

To fully leverage the RNA-sequencing data in this study, we performed pathway enrichment analysis in order to identify the immunological and cellular pathways which show significant enrichment for genes significantly modulated between two groups or which significantly correlated with disease severity. For this, we used an application called ClueGO in Cytoscape (33). This function allows the user to input a list of significantly modulated genes and then uses Fisher’s exact testing to identify the pathways which show statistically significantly higher enrichment for genes from the input list than would be expected at random from the genome. Pathways were taken from the Gene Ontology (GO) (34), Kyoto Encyclopedia of Genes and Genomes (KEGG) (35) and Reactome databases (36). Then an additional step is taken to produce a network-based analysis by using kappa statistics to identify pathways have significant overlap in enriched genes and then grouping these together into distinct functional nodes (color coded individually) to identify groups of pathways with similar and/or redundant function. To accomplish this, ClueGO using a matrix of genes with associated terms (pathways) and calculates a similarity matrix using kappa statistics to identify terms (pathways) which shows the strength of association between terms (pathways) based on containing the same enriched genes. The kappa score can be tuned from 0 to 1 to increase or restrict network connectivity as needed. Consolidating redundant pathways into nodes provides easier network visualizations to demonstrate multiple key changes in immunology and/or cellular function. We utilize this powerful and open-source tool in order to dig into our transcriptional data and identify the main immunological impacts of changes in the circulating lipid profile and transcriptional profile from individuals with differing degrees of COVID-19 disease severity.

## Results

### Increased expression of genes involved in the synthesis of PUFAs-derived lipid mediators is increased in the nasopharyngeal mucosa of patients with high WHO score

We re-analyzed data from nasopharyngeal (NP) swabs collected from 58 individuals from the University of Mississippi Medical Center (UMMC) between April and September 2020. Full cohort details can be found in a previous publication (24). Quantitative PCR analysis of NP swabs in this patient’s cohort revealed that arachidonate 5-lipozygenase 5 (*alox5*) a gene that utilize PUFAs a substrate to generate leukotriene, a family of eicosanoid inflammatory mediators, is over-represented in mast cells (Fig. 1A). However, recent studies show that *alox5* plays also an active role in the synthesis of specialized pro-resolving mediators (SPMs) a large class of signaling molecules implicated in the resolution of inflammation (37). *Alox15*, another arachidonate 5-lipozygenase enzyme is widely distributed in most cell types of the naso-pharynx, including ciliated cells, developing secretory and goblet cells and goblet cells but excluded form mast cells (Fig. 1A). Both *alox15* and *alox15B* play a major role in the synthesis of the SPMS lipoxins and resolvins. Specifically, violin plot analysis revealed that *alox15* is predominantly expressed in COVID-19 patients with high WHO score (6–8) in developing ciliated cells, goblet cells, deuterosomal cells, Ionocytes and developing secretory and goblet cells (Fig. 1B). Confirming the implication of SPMs synthesizing gene expression in COVID-19 patients with high WHO score, we detected a high expression in goblet cells of phospholipase A2 group IVA (*pla2G4a*), a gene implicated in the hydrolysis of membrane phospholipids to release arachidonic acid to be subsequently metabolized in eicosanoids (Fig. 1C); *alox15B* and *aloxE3*, another lipoxygenase enzyme that in humans form a cluster of enzymes with *alox12* and *alox15B* on chromosome 17. These results may implicate that upon severe COVID-19 infection, developing and secretory goblet cells and mature goblet cells will actively synthesized SPMs as a mean of fighting the inflammation resulting from SARS-CoV-2 infection.

**Figure 1:**
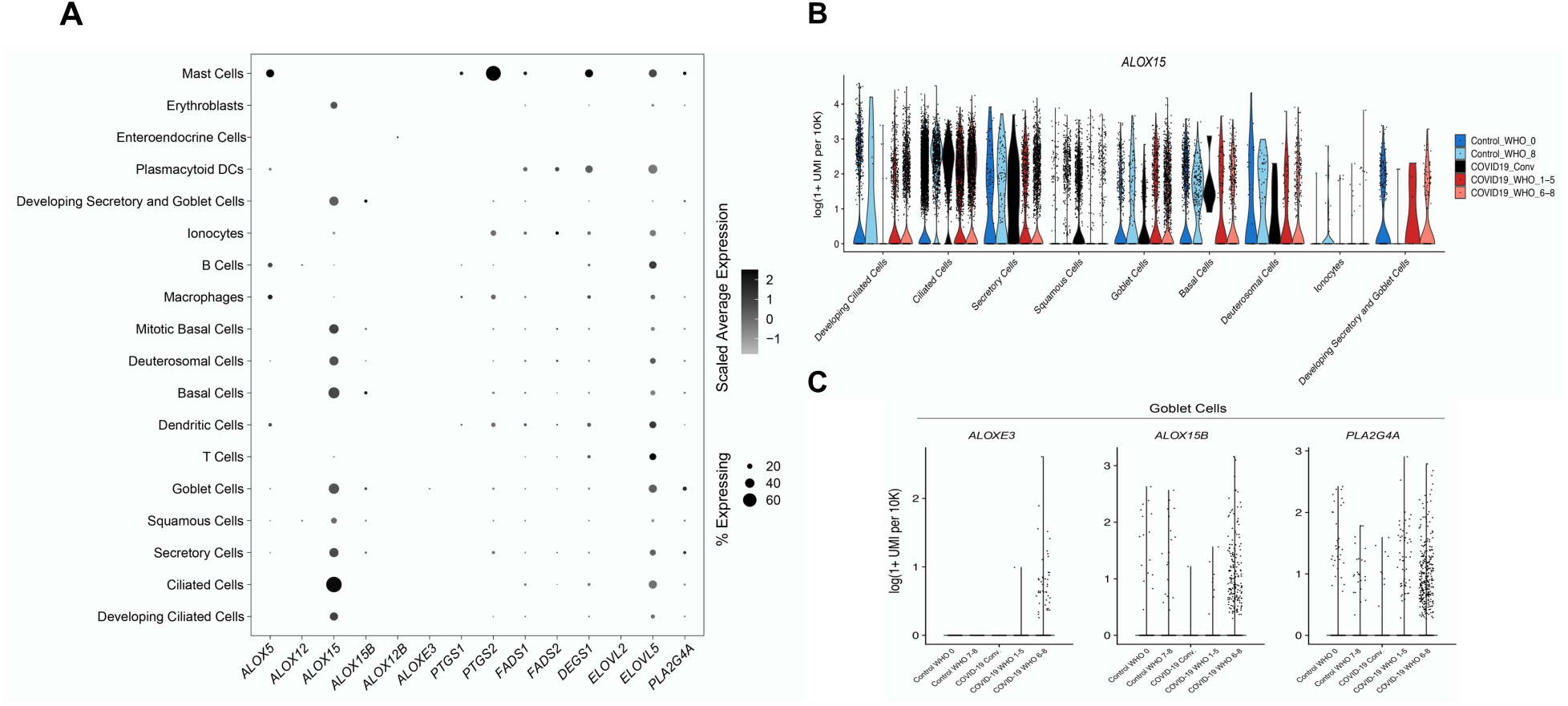
Expression of resolving synthesizing enzymes in covid-19 patients. (A) Expression of alox enzymes and other fatty acid related genes. Dot size represent fraction of cell type (rows) expressing a given gene (columns). Dot hue represent scaled average expression by gene column. (B) Select differentially expressed genes between control non-SARS-CoV-2 cells and SARS-CoV-2 RNA+ cells from participants with control WHO score of 0 in dark blue; control WHO score of 0 in light blue; COVID-19 convalescent patients in black; mild or moderate COVID-19 in red and severe COVID-19 in pink. (C) Violin plots of selected genes among goblet cells in Control WHO 0; Control WHO 7-8; COVID-19 convalescent patients; COVID-19 WHO 1-5 and COVID-19 WHO 6-8.

### Eicosanoid precursors demarcate patients with more severe COVID-19 disease from those with less severe disease

To understand better why SPMs synthesizing gene expression is increased COVID-19 patients with high WHO score we utilize a subset of 20 individuals from the previous cohort for which we had peripheral blood mononuclear cell (PBMC) samples as described in the methods section. Using this cohort, we tried to understand how circulating lipid metabolites are modulated in SARS-CoV-2 infection. We compared the lipid profiles of healthy controls with hospitalized patients with SARS-CoV-2. Using Principal Component Analysis (PCA) to visualize the overall variance in our dataset, we did not observe any difference in clustering of samples between healthy controls and those with SARS-CoV-2 (Supplemental Figure 1). A row normalized heatmap was used to visualize the 31 lipid metabolites measured (Fig. 2A). Univariate comparison of lipid metabolites using Mann-Whitney U-test revealed that 6 lipid metabolites were significantly (p≤0.05) increased in SARS-CoV-2 patients compared to healthy controls (red asterisks in Fig. 2A). These include the inflammatory mediators Arachidonic Acid (AA) and Eicosapentaenoic Acid (EPA) which are known to give rise to eicosanoids and may serve as drivers of systemic inflammation seen in SARS-CoV-2. Further inspection of the heatmap reveals that there are two potential clusters of SARS-CoV-2 patients, one exhibiting general elevated levels of lipid mediators (boxed in black) compared to another cluster with low expression of lipids even compared to controls. Of interest, those with higher levels of lipids have higher WHO score. PCA analysis of just the SARS-CoV-2 infected samples confirms those with a WHO score <5 separate on PC1 and PC2 from those with a WHO Score >5 (Fig. 2B). To explore this heterogeneity in lipid profiles between those with more severe disease scores, we again performed univariate analysis between the two groups using Mann-Whitney U-tests (Fig. 2C). We identify that some lipid metabolites identified as heightened in SARS-CoV-2 (AA, Docosahexaenoic acid (DHA), Docosapentaenoic acid (DPA), and Docosatetraenoic acid) are also increased significantly in patient samples with more severe SARS-CoV-2 disease. Notably, additional inflammatory lipids including Eicosatrienoic acid and Eicosadienoic acid (metabolites leading to AA and Eicosanoids) and Stearic acid were also significantly elevated in samples from patients with more severe disease. These data highlight that individuals with inflammatory lipid profiles are more likely to have a more severe disease score. To confirm this, we performed spearman correlations of lipid metabolites with WHO score (Fig. 2D). Consistent with our prior findings, key inflammatory lipids including AA (rho=0.742, p=0.018) and EPA (rho=0.736, p=0.019) along with the fatty acids DHA (rho=0.736, p=0.019) and DPA (rho=0.699, p=0.029) were significant positive correlates of WHO disease severity score. Lineolic acid (rho=0.0644, p=0.05) and its related metabolites (a-linoleic acid [rho=0.663, p=0.042] and Dihomo-y-linolenic acid [0.657, p=0.045]) which are precursors for Eicosanoids were also identified as positive correlates of disease severity. Combined, these data clearly illuminate that individuals with heightened circulating lipids involved in inflammatory responses through the generation of eicosanoids have more severe disease providing a direct link between inflammatory lipids and the spread of inflammatory disease during SARS-CoV-2.

**Figure 2:**
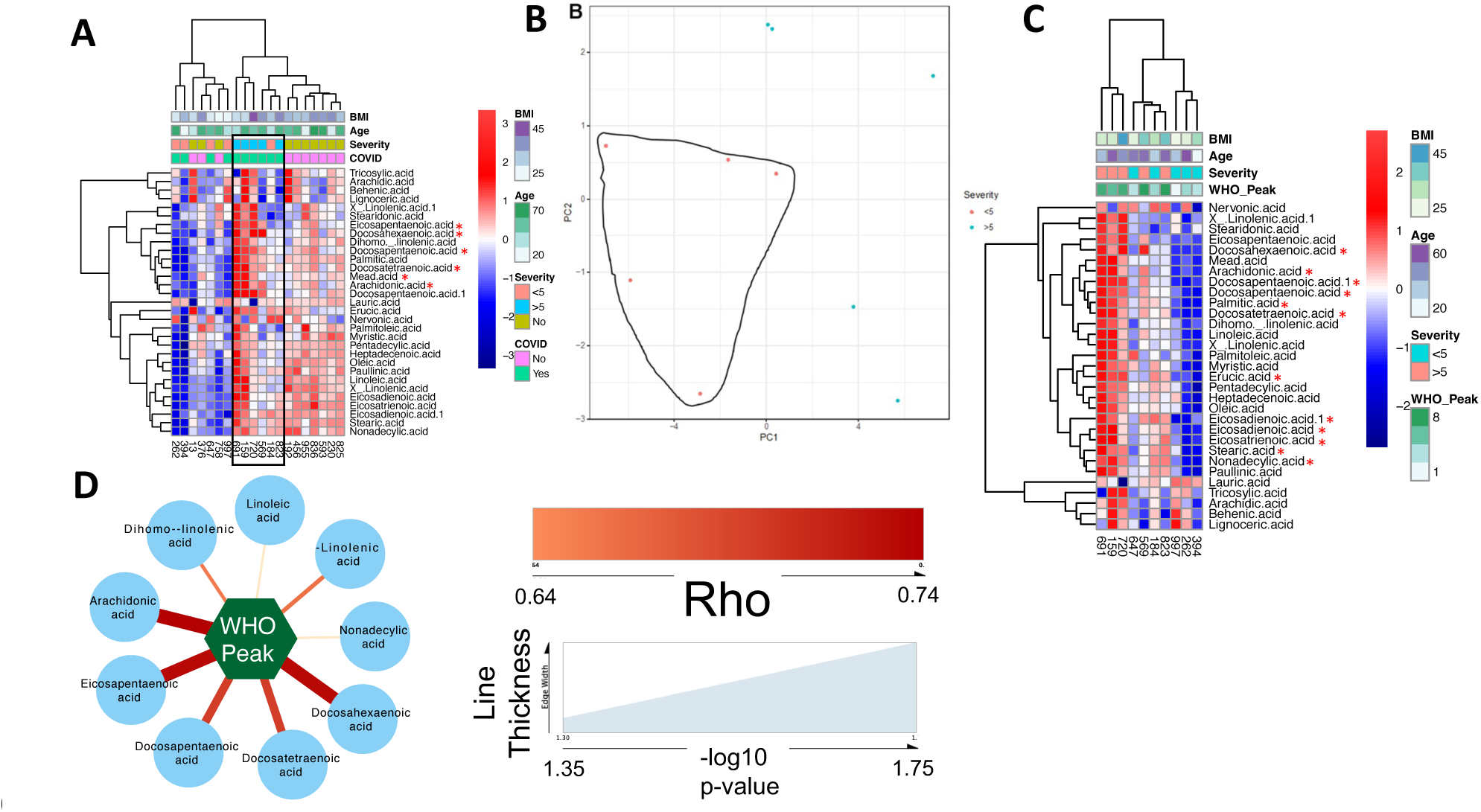
Arachidonic acid and eicosanoid metabolites are significantly increased in COVID-19 and correlated with disease severity. (A) Row normalized heatmap of lipidomic expression profiles comparing hospitalized individuals with COVID-19 (n=10) and health controls (n=10). *=p<0.05 by Mann-Whitney U-test. (B) PCA analysis showing that participants with a high disease severity (WHO Score >5) separate from those with low disease severity (WHO Score <5). (C) Row normalized heatmap visually confirms distinct metabolic profiles associated with heightened disease severity. *=p<0.05 by Mann-Whitney U-test. (D) Network visual of Spearman correlations of lipid metabolites with WHO Score, identifying arachidonic acid and eicosanoid metabolites as key lipid biomarkers of disease severity. Color of line=rho; Thickness of line= negative log10 of p-value

### Pathways of cell cycle progression, DNA damage and innate immune function are differentially modulated transcriptionally between patients with high and low WHO disease scores

Having established a unique lipid profile exists between patients with high vs. low WHO disease score, we next wanted to investigate how transcriptional responses are differentially modulated between these two groups. Using normalized read counts from each sample, we first performed an unsupervised analysis of variance of the data using PCA. Using this analysis, we observe two main clusters of samples with one comprised of 3/5 patients with low WHO score and the other with 3/4 patients with high WHO score (Fig. 3A). A heatmap plotting the top 100 DEGs confirms that there are distinct transcriptional profiles between patients with low and high WHO disease score severity (Fig. 3B). Using unbiased clustering by Euclidian distance, we observe that the two groups of patients separate on this heatmap providing clear evidence that these groups have distinct transcriptional profiles. The genes which were significantly induced and reduced (adjusted p≤0.05) in the WHO high score patients are represented in separate network models (Supplemental Fig. 2A,B). To better understand the biology underlying these differences in transcriptional responses, we performed pathway enrichment analysis on genes which were significantly (p<0.05) modulated between the two groups using the ClueGO app in Cytoscape and pathways from the Gene Ontology, KEGG and Reactome datasets. Separate analyses were performed for genes significantly induced in high WHO score and genes significantly reduced in high WHO score. Our analysis revealed no immune pathways which were significantly (p≤0.05) enriched for genes which were induced in WHO high score patients (Supp. Fig. 2C) though protein trafficking and response to hypoxia, likely due to pathologically low levels of O2 saturation, were increased in high disease severity. We identified several key pathways and nodes which were enriched for genes which were significantly reduced in WHO score high patients. These include pathways related to innate immunity, interactions between lymphoid and non-lymphoid cells, Interleukin signaling, and RHO GTPase function (Fig. 3C). Although inflammation and innate immune activation are a hallmark of severe COVID-19 disease, these data suggest the potential that within those who have severe disease there is impaired innate immune function in those with the most severe disease and that this may be a contributing factor to their poor disease outcomes. Pathways induced in WHO high score were related to Golgi-ER trafficking, protein SUMOylation, and cellular response to hypoxia, likely reflective of reduced respiratory function and blood oxygen levels seen with increasing disease severity (Supplementary Fig. 2C). We next correlated gene expression with WHO score across the entire cohort of 10 patients with sequencing and performed pathway enrichment analysis using ClueGO as before. Genes which are significant and positive correlates of WHO score (adjusted p≤0.10) were enriched in pathways of cell cycling (G1/S checkpoints, G2/M transition) and homologous recombination in response to DNA damage indicating those with high disease score undergo augmented cell cycling and DNA damage (Fig. 4A). This is consistent with a previous report demonstrating that cell cycle dysregulations are common changes in severe COVID-19, suggesting an association with disease severity (38). We also observed an increase in pathways of the unfolded protein response and protein targeting to the ER suggesting heightened ER stress in these patients which may also contribute to inflammation and pathology. When we look at pathways enriched for genes which significantly and negatively correlate with WHO score reduced genes (adjusted p≤0.10), we identify pathways and nodes associated to IL-1 and inflammasome function as well as IFN signaling (Fig. 4B). If we perform pathway analysis on genes which correlate with a rho>0.80 or <-0.80 and adjusted p≤0.15 we again see significant enrichment in pathways of cell cycling among positively correlating genes and pathways of innate immunity as enriched for negatively correlating genes (Supplemental Fig. 3A-3B). These findings support an association between lower innate immune activation (seen in Fig. 3C) in those with high WHO disease score and further highlight that there may be critical differences in the regulation of innate immune function between SARS-CoV-2 infected patients with a high vs a low WHO disease severity score. Further investigation of these differences could be critical for understanding how to treat patients who present with more severe COVID-19 disease.

**Figure 3:**
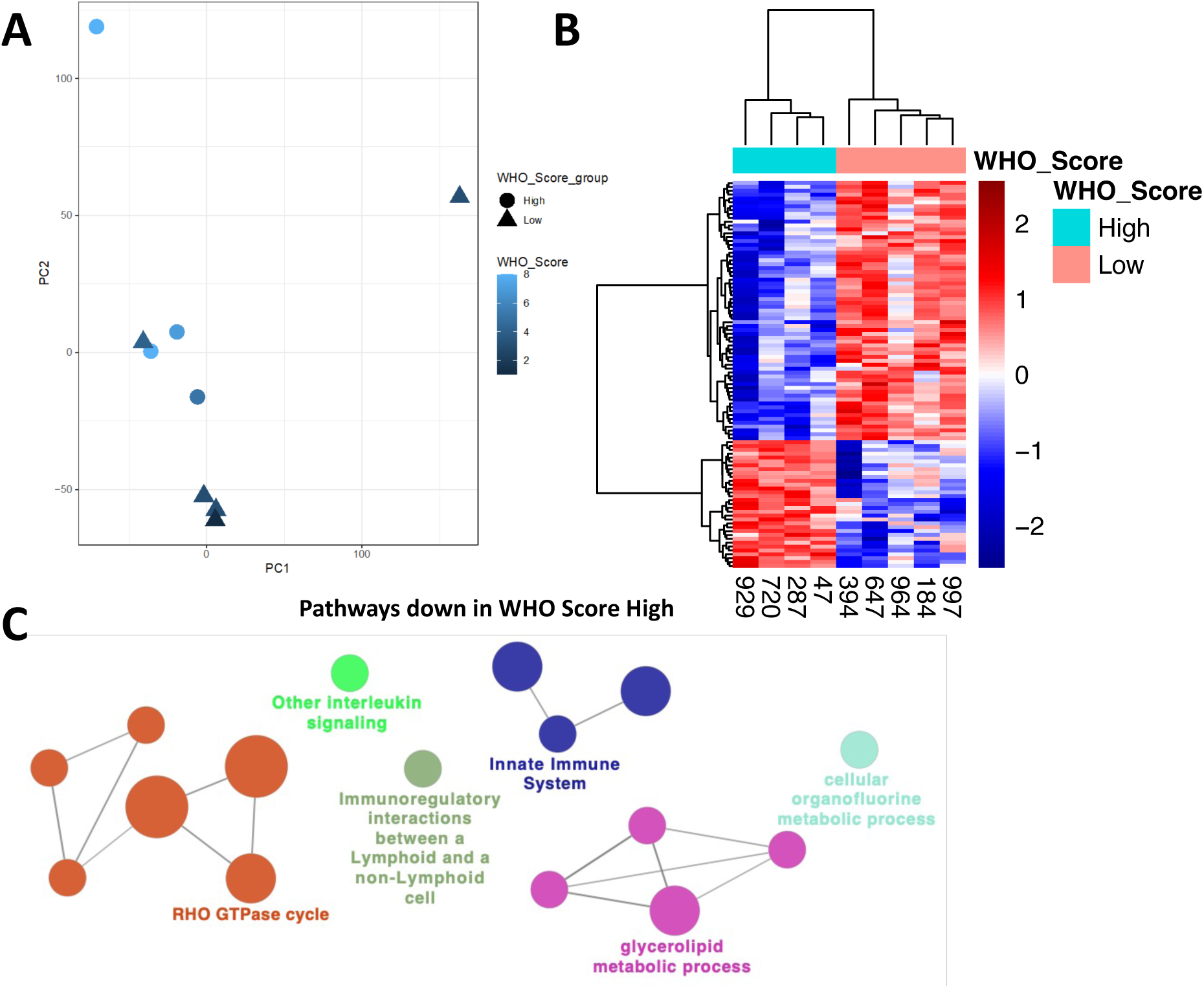
High disease severity in COVID-19 is associated with reduced expression of genes in pathways of innate immunity and cytokine signaling. RNA-sequencing was performed on n=9 participants with COVID-19. (A) PCA analysis reveals that the transcriptional profile of 4 of 5 individuals with high WHO disease severity separate from the transcriptional profile of those with low WHO disease severity. (B) Row normalized heatmap of top 100 differentially expressed genes showing clear differential expression of these genes between the low and high WHO disease severity groups. (C) Network-based pathway enrichment analysis of DEGs (adjusted p<0.05) between low and high disease severity score groups using ClueGO in Cytoscape. Genes with reduced expression in high disease severity are significantly (p<0.05 by Fisher’s exact-test) enriched in pathways of innate immunity, cytokine signaling, immune cell interaction, GTPase function and lipid metabolism.

**Figure 4:**
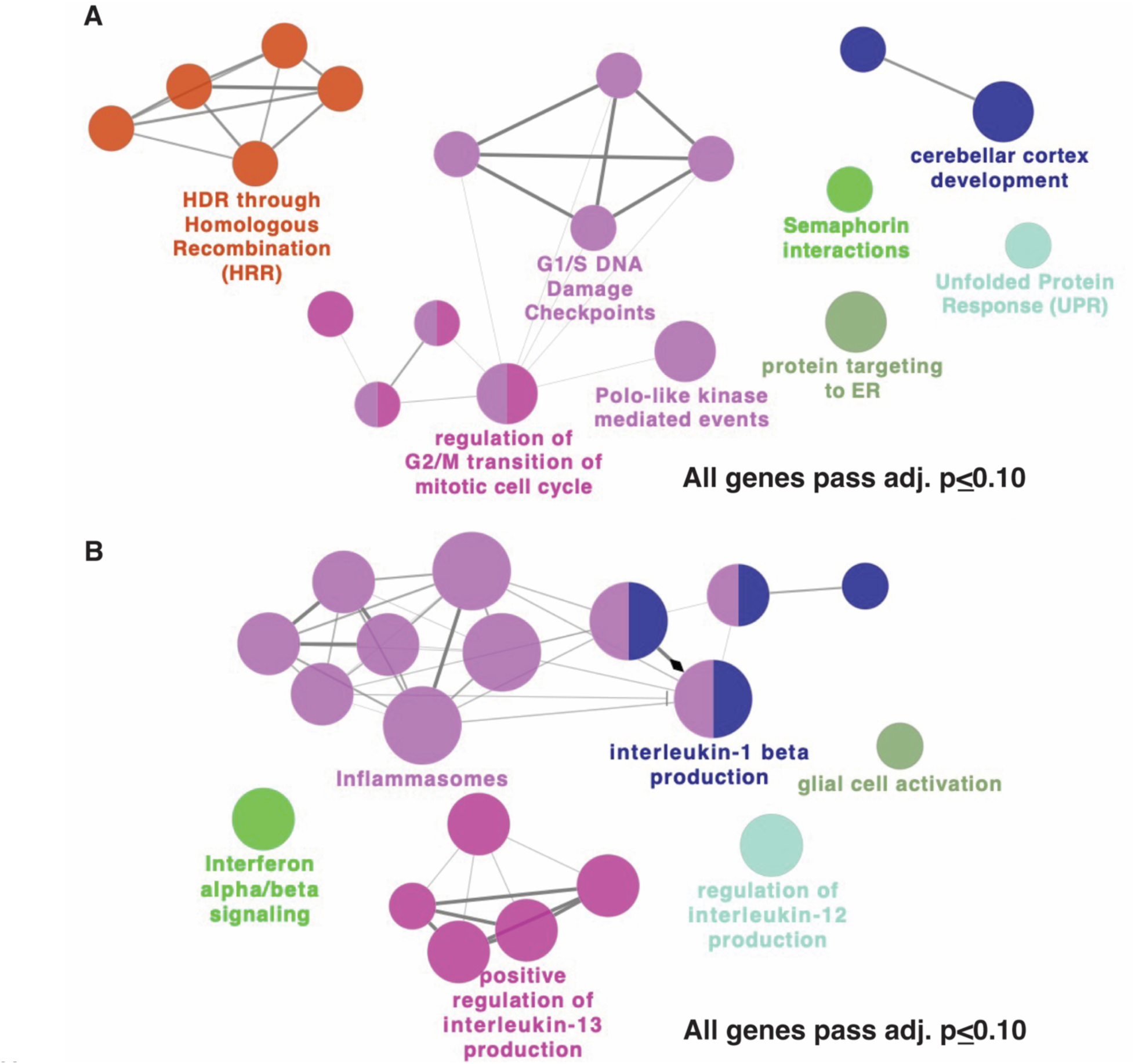
Pathways of cell cycling and DNA damage correlate positively with COVID-19 disease severity. Spearman correlations were performed between gene expression and WHO disease severity score. Significant (adjusted p≤0.10) positively (A) and negatively (B) correlating genes are mapped to enriched pathways using ClueGO. (A) Pathways of cell cycling, DNA damage DNA repair, and ER stress are positively correlated with disease severity. (B) Pathways of Interleukin-1/Inflammasomes, Interferons, and IL-13 cytokine production are enriched for genes which are negative correlates of COVID-19 disease severity.

### Transcriptional signatures of COVID-19 disease are marked by heightened pathways of cell cycling and reduced pathways of antigen presentation and response to infectious diseases

To determine if the changes in transcriptional signatures we observed were specific to participants with WHO high score or a feature of all COVID-19 samples elevated in high WHO score, we compared our RNA-sequencing in the 10 COVID-19 participants with 10 healthy controls. We identified 2021 genes which were significantly modulated between COVID-19 samples and healthy controls at a nominal p-value of 0.05, with 101 of these passing correction for multiple testing (adj. p <0.05) (volcano plot in Fig. 5A). Of those 101, 24 were increased in COVID-19 samples while 77 were increased in healthy control samples. A row normalized heatmap shows the consistent and clear dichotomy in expression of these two sets of genes between COVID-19 and control samples (Fig. 5B). Among the genes induced in COVID-19 is CCR2, a chemokine receptor which binds to MCP-1 to mediate myeloid chemotaxis and inflammation (Supplemental Fig. 4A). JAK1, a kinase involved in cytokine signaling including interferon signaling, and NFATC1, a key transcription factor in T cell signaling, along with Class II MHC genes (HLA-DPA1, HLA-DMB, and HLA-DOA) were reduced in COVID-19 samples indicating a potential perturbation in IFN signaling as well as CD4+ T cell priming and activation (Supplemental Fig. 4B-D). Pathway enrichment of the significantly modulated genes using the Reactome database reveals that genes induced in COVID-19 are part of pathways involved in amino acid metabolism, cell cycling, and G-protein receptor signaling (Fig. 5C). Multiple immune pathways including MHC Class II presentation, cytokine signaling and infectious disease responses are reduced in COVID-19 samples (Fig. 5D). Pathways of RNA processing, protein translation, metabolism and DNA damage repair are also reduced suggesting global defects in cellular function are associated with COVID-19 in this cohort. These data show that increases in cell cycling and reductions in innate immunity are general markers of COVID-19 as well as potential drivers of disease severity, with higher cell cycling and lower innate immune function in participants with more severe disease.

**Figure 5:**
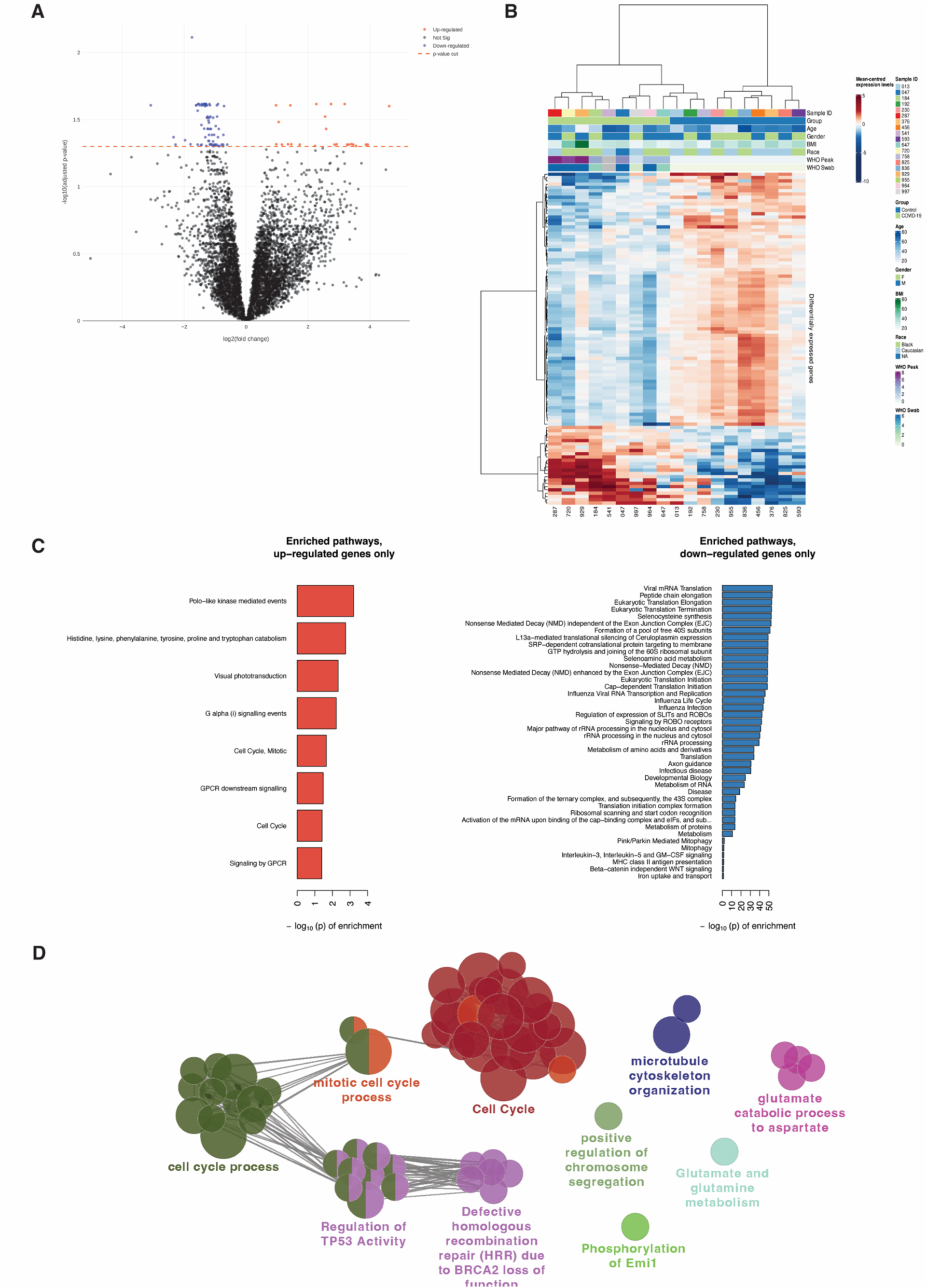
Cell cycling and GPCR signaling pathways are significantly increased in patients with COVID-19 compared to healthy controls. RNA-sequencing analysis comparing patients with COVID-19 (n=9) and healthy controls (n=10). (A) Volcano plot showing increased and decreased expression of genes. Dots above the line are significantly (p<0.05). (B) Row normalized heatmap of top 100 DEGs showing distinct transcriptional profiles between healthy individuals and those with COVID-19. (C) Pathway enrichment analysis by GSEA identified pathways of amino acid metabolism, cell cycling, and GPCR signaling being significantly enriched for genes increased in COVID-19. Pathways enriched for genes down-regulated in COVID-19 disease include response to viral infections, cytokine signaling, antigen processing/presentation, and protein translation. All pathways pass fdr cutoff of <0.05. (D) Network-based pathway enrichment by ClueGO confirms that expression of genes in pathways of cell cycling and amino acid metabolism is increased in COVID-19 disease.

### The inflammatory lipid Eicosapentaenoic acid correlates with heightened cell cycling and DNA damage pathways observed in patients with high WHO score

Having provided foundational evidence for a critical role of circulating inflammatory lipids in modulating disease severity and identifying distinct transcriptional profiles associated to high and low WHO disease score, we next investigated whether a prototypical inflammatory lipid EPA which is a known precursor to eicosanoids was correlated with transcriptional changes observed in our cohort. Using the subjects in common to lipid metabolomics and transcriptomics (n=5), we performed spearman correlations to identify genes which positively and negatively correlated with levels of EPA. We identified 88 genes which were significant positive correlates (p≤0.05) of EPA which we have visualized in a network model (Supplemental Fig. 5A). Interestingly, we found no genes whose expression correlated significantly and negatively (p≤0.05) with EPA. Pathway enrichment analysis using ClueGO reveals that these 88 genes map to pathways of mitotic centrosomes and centrioles, N-Glycan biosynthesis, and ECM interactions (Supplemental Fig. 5B). Given the low number of patients (n=5) in this analysis and to expand on the identification of pathways involved in cell cycling and mitosis, we performed a second pathway enrichment analysis in ClueGO using any gene which positively correlated with a p-value of ≤0.10 (rho≥0.872). This analysis identified multiple pathways of cell cycle, mitosis, microtubule and chromosome organization, and DNA damage/TP53 signaling as being enriched for genes which positively correlate with EPA (Fig. 5E). Only 4 genes were found to correlate negatively with a p-value of ≤0.10 and these were HDAC2, AFTPH, ARMC6, and KIAA1549. HDAC2 is a potent regulator of transcriptional silencing by removing acetylation marks from histones, leading to epigenetic silencing of target loci. Further investigation can be performed to determine if there are any changes in histone acetylation and chromatin modification which are associated to higher levels of inflammatory lipids. Altogether, these data provide the first direct link between a circulating eicosanoids lipid profile and changes in transcriptional regulation which promote enhanced cell cycling, mitosis, and DNA damage while potentially modulating the epigenetic landscape of circulating immune cells. These factors may contribute to rapid cell turnover and an inability to maintain normally homeostatic proliferation in the blood of patients with severe COVID-19 disease.

## Discussion

While the deployment is widely available, SARS-CoV-2 vaccines has resulted in a significant reduction in the levels of hospitalized and severe COVID-19 following infection, it is still important to better understand the mechanisms which underly progression to severe disease to develop more effective therapies to prevent morbidity and mortality associated with severe disease. Multi-omics approach has been instrumental to characterize changes in cellular and metabolic pathways that could be associated to disease severity. Among the most frequently reported findings is aberrant immune cell activation (39,40), and lipid metabolism impairment (41). Consistently, lipid panel analysis by other groups have highlighted the role of lipid metabolism and inflammatory lipid mediators in severe disease progression, identifying changes in SPMs in serum and plasma (21,42) and lungs (43). By integrating lipidomic analysis with transcriptomics we aim to contribute to further understand the immunopathology underlying severe COVID-19.

Our initial results from transcriptomics analysis obtained using the patient cohort presented previously (24), showed that several genes implicated in the synthesis specialized pro-resolving mediators (SPMs) such as *alox5* and *alox15* are highly expressed in nasopharyngeal swabs from COVID-19 patients. These alox genes encode enzymes that synthesize lipoxins, the lead family of pro-resolving mediators, from AA, EPA or DHA (44). Elevated ALOX5 activity in the circulation has been found in patients with severe COVID-19, suggesting an association between this enzyme’s metabolic products and severity progression during SARS-Cov-2 infection (45). High WHO score COVID-19 patients display an increase in *alox15* expression as one would expect when the organism has to resolve an inflammation. However, it is striking to observed that the highest levels of *alox15* expression in the ciliated cells, secretory cells and squamous cells of the nasopharynx is detected in covid-19 patients in the convalescent phase, indicating that high level of production of lipoxins and SPMs is required to clear the organism from the COVID-19 induced inflammation. This observation that lipoxins and SPMs play a pivotal role in the recovery from COVID-19 infection is substantiated by a recent report linking disease severity to plasma levels of SPMs (42). Palmas et al. established a correlation between peripheral blood SPMs concentration and patient outcome. Patients who did not survive the COVID-19 infection were those with the lowest concentration of SPMs in their peripheral blood (42). Our results concurred with the observation made previously that synthesis of SPMs or levels of circulating SPMs may be linked with patient survivor following COVID-19 infection. However, it has to be noted that the increase in alox genes expression was mostly detected in NP swabs analysis and not PBMCs highlighting that these two cells population might respond differently to COVID-19 infection and recovery.

Comparison of the intracellular lipid profiles in PBMCs of participants with severe COVID-19 to those of healthy controls identified 6 lipids which were significantly elevated in COVID-19 samples including lipid metabolites which given rise to Eicosanoids, key mediators of inflammation in response to injury and infection. The heatmap revealed two distinct clusters of COVID-19 samples, one with elevated levels of lipids compared to controls and one with decreased lipid levels compared to controls. Interestingly, those with enhanced lipid profiles had higher disease severity (>5) as measured by WHO score, a finding corroborated by unbiased PCA which revealed samples from WHO score >5 separated from those with lower disease score. When we only compare lipids within COVID-19 samples, we observed that 12 lipids including multiple involved in Eicosanoid synthesis are elevated in COVID-19 disease and in fact that these lipids are significant positive correlates of disease severity measures. Since these lipids are measured intracellularly in PBMCs, this study provides the first clear evidence that PBMCs can serve as a major source of circulating inflammatory lipids which can contribute to systemic inflammation and disease pathology.

Comparing the transcriptional responses in WHO score high vs. WHO score low participants revealed further dichotomy of these two populations. Interestingly, pathways associated with innate immune function were reduced in participants with a high WHO score suggesting that even those these individuals have prolonged inflammation due to SARS-CoV-2 infection their innate immune system may not be able to control infection as efficiently, potentially contributing to heightened disease pathology. Cellular response to hypoxia was one of the pathways increased in samples from WHO high score patients. This is consistent with the clinical outcomes of these individuals who presented poor respiratory function, showing this is not limited to tissues but also spreads to circulating immune cells.

To provide more granularity to the relationship between transcriptional changes and disease severity, we identified the genes which were significantly modulated with an adjusted p-value of 0.10 and then performed pathway enrichment. This analysis validated that heightened cell cycling and DNA damage were positive correlates of disease severity while reductions in innate immunity, IL-1, Interferon and viral sensing pathways were correlated with lower disease severity. Our data provides greater clarity into the systemic activation of the immune function across the clinical manifestation spectrum, suggesting that while a global look at disease severity finds an association with inflammation, those with the most severe disease have reduced inflammation which is reflective of more dysfunctional innate immune responses that are incapable of controlling the viral infection as efficiently.

In summary, we were able to show that pathways involved in cell cycling, along with GPCR signaling, are global features of COVID-19 disease. Reductions in pathways of innate immune function were also observed in all COVID-19 samples compared to controls. The magnitude of these changes in expression are associated with augmented disease severity. Our Multi-Omics approach aims to provide a new level of granularity to our understanding of potential drivers of severe COVID-19. By identifying inflammatory lipids and their association to heightened cell cycling and reduced innate immune function during severe COVID-19, we identified multiple potential therapeutic targets along the cascade of immunopathology, from some of the earliest inflammatory mediators such as Eicosanoids, to their downstream cytokines/innate immune pathways. Enhancing our understanding of the mechanisms involved in disease severity can have profound implications in the discovery of novel interventions to manage severe cases of COVID-19 and reduce the burden of this virus.

## Data Availability

All data produced in the present work are contained in the manuscript.

## Acknowledgements

The authors would like to thank the patients and their families for participating in this study. We also wish to thank the UMMC staff for their crucial support to make this research possible.

## Author contributions

Y.G. and S.C.G. directed the study. J.A.T., S.C.G., M.G.P. and Y.G. wrote the manuscript. A.O., C.G.K.Z., T.G.W., H.L., H.B.W., N.S.G. and A.K.S recruited patients and collected samples. A.O., T.O.R, B.H.H. and J.O-M. performed the experiments. J.A.T. and Y.G. analyzed the data. All authors provided input into data analysis and approved the final version of the manuscript.

## Supplementary Table and Figure Legends

**Supplemental Table 1:** Participant characteristic nasopharyngeal covid-19 cohort.

|  | Control (WHO score 0) | Intubated Control (WHO score 7-8) | COVID-19 Mild/Mod. (WHO score 1-5) | COVID-19 Severe (WHO score 6-8) | COVID-19 Conv. (WHO score 0) |
| --- | --- | --- | --- | --- | --- |
| Case Number | 25.9% (15/58) | 10.3% (6/58) | 24.1% (14/58) | 36.2% (21/58) | 3.4% (2/58) |
| Sex |  |  |  |  |  |
| Male | 40% (6/15) | 83.3% (5/6) | 57.1% (8/14) | 52.4% (11/21) | 50% (1/2) |
| Female | 60% (9/15) | 16.7% (1/6) | 42.9% (6/14) | 47.6% (10/21) | 50% (1/2) |
| Age (years) |  |  |  |  |  |
| Minimum | 27 | 33 | 19 | 28 | 20 |
| Median (IQR) | 58 (16) | 65.5 (31) | 49.5 (17.8) | 62 (13) | N/A |
| Maximum | 73 | 71 | 69 | 84 | 57 |
| Race |  |  |  |  |  |
| Black/African American | 66.7% (10/15) | 66.7% (4/6) | 71.4% (10/14) | 61.9% (13/21) | 50% (1/2) |
| White | 33.3% (5/15) | 33.3% (2/6) | 28.6% (4/14) | 23.8% (5/21) | 50% (1/2) |
| American Indian | 0% (0/15) | 0% (0/6) | 0% (0/14) | 14.3% (3/21) | 0% (0/2) |
| Ethnicity |  |  |  |  |  |
| Hispanic | 0% (0/15) | 0% (0/6) | 0% (0/14) | 4.8% (1/21) | 0% (0/2) |
| Not Hispanic | 100% (15/15) | 100% (6/6) | 100% (14/14) | 95.2% (20/21) | 100% (2/2) |
| BMI |  |  |  |  |  |
| Median (IQR) | 37.5 (14.4) | 30.5 (18.1) | 23.0 (11.6) | 31.9 (14.2) | 40.7 |
| Pre-Existing Conditions |  |  |  |  |  |
| Diabetes | 40% (6/15) | 33.3% (2/6) | 28.6% (4/14) | 71.4% (15/21) | 0% (0/2) |
| Chronic Kidney Disease | 6.7% (1/15) | 0% (0/6) | 7.1% (1/14) | 19.0% (4/21) | 0% (0/2) |
| Congestive Heart Failure | 6.7% (1/15) | 16.7% (1/6) | 0% (0/14) | 4.8% (1/21) | 0% (0/2) |
| Lung Disorder | 6.7% (1/15) | 16.7% (1/6) | 28.6% (4/14) | 38.1% (8/21) | 0% (0/2) |
| Hypertension* | 86.7% (13/15) | 50% (3/6) | 42.9% (6/14) | 81.0% (17/21) | 0% (0/2) |
| Inflammatory Bowel Disease | 13.3% (2/15) | 0% (0/6) | 0% (0/14) | 0% (0/21) | 50% (1/2) |
| Treatment |  |  |  |  |  |
| Corticosteroids | N/A | 33.3% (2/6) | 42.9% (6/14) | 66.7% (14/21) | N/A |
| Remdesivir | N/A | 0% (0/6) | 14.3% (2/14) | 4.8% (1/21) | N/A |
| 28-Day Mortality*** | 0% (0/15) | 33.3% (2/6) | 0% (0/14) | 76.2% (16/21) | 0% (0/2) |
| Experiment Performed |  |  |  |  |  |

**Supplemental Figure 1:**
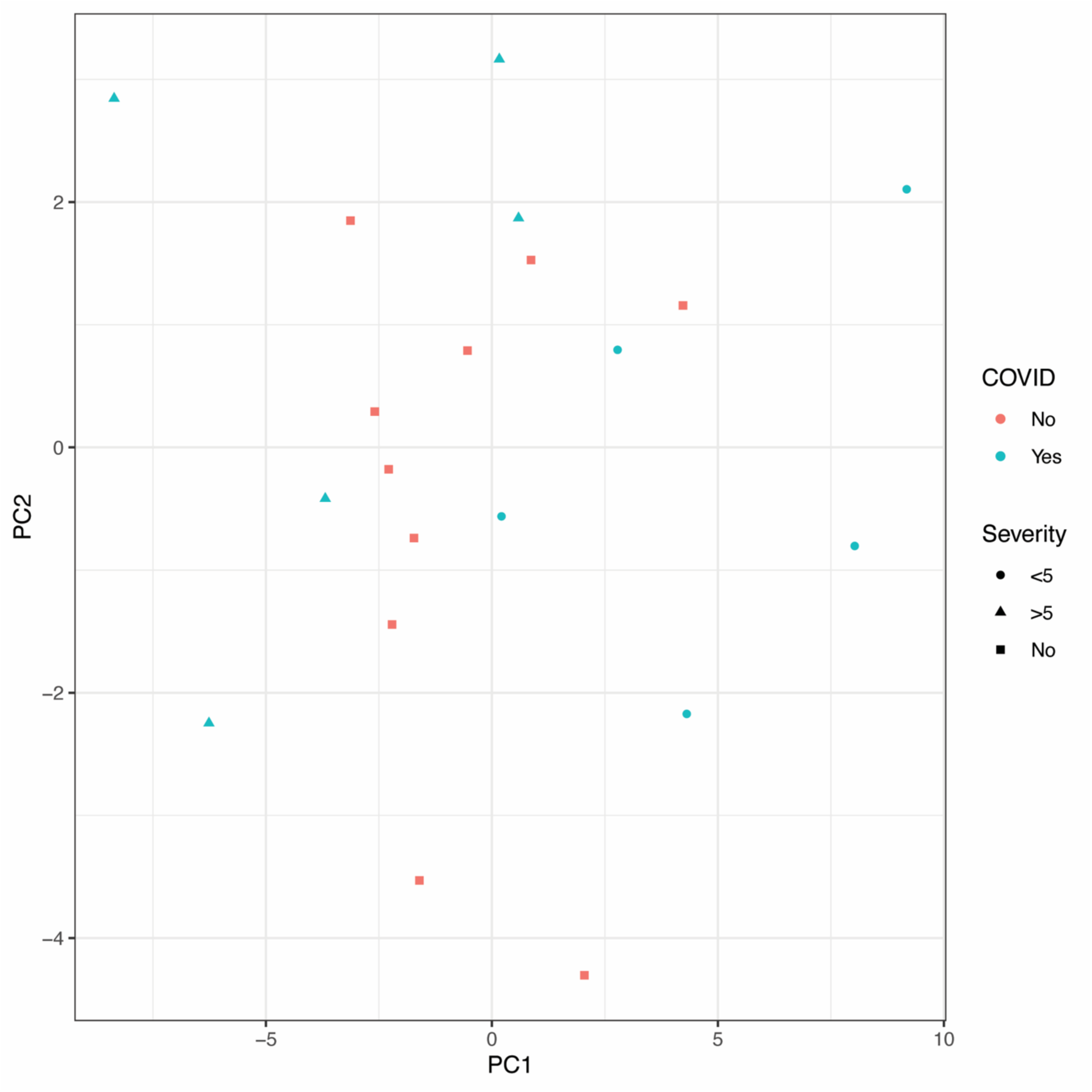
Lipidomic profile of healthy controls falls between two clusters of COVID-19 patients. PCA analysis of lipidomic profiles comparing healthy control (red) with participants with COVID-19 (green). Healthy controls do not cluster distinctly from COVID-19 patient samples. COVID-19 high disease severity samples (circles) separate from COVID-19 low disease severity samples (triangles) with healthy controls falling in the middle of the two groups.

**Supplemental Figure 2:**
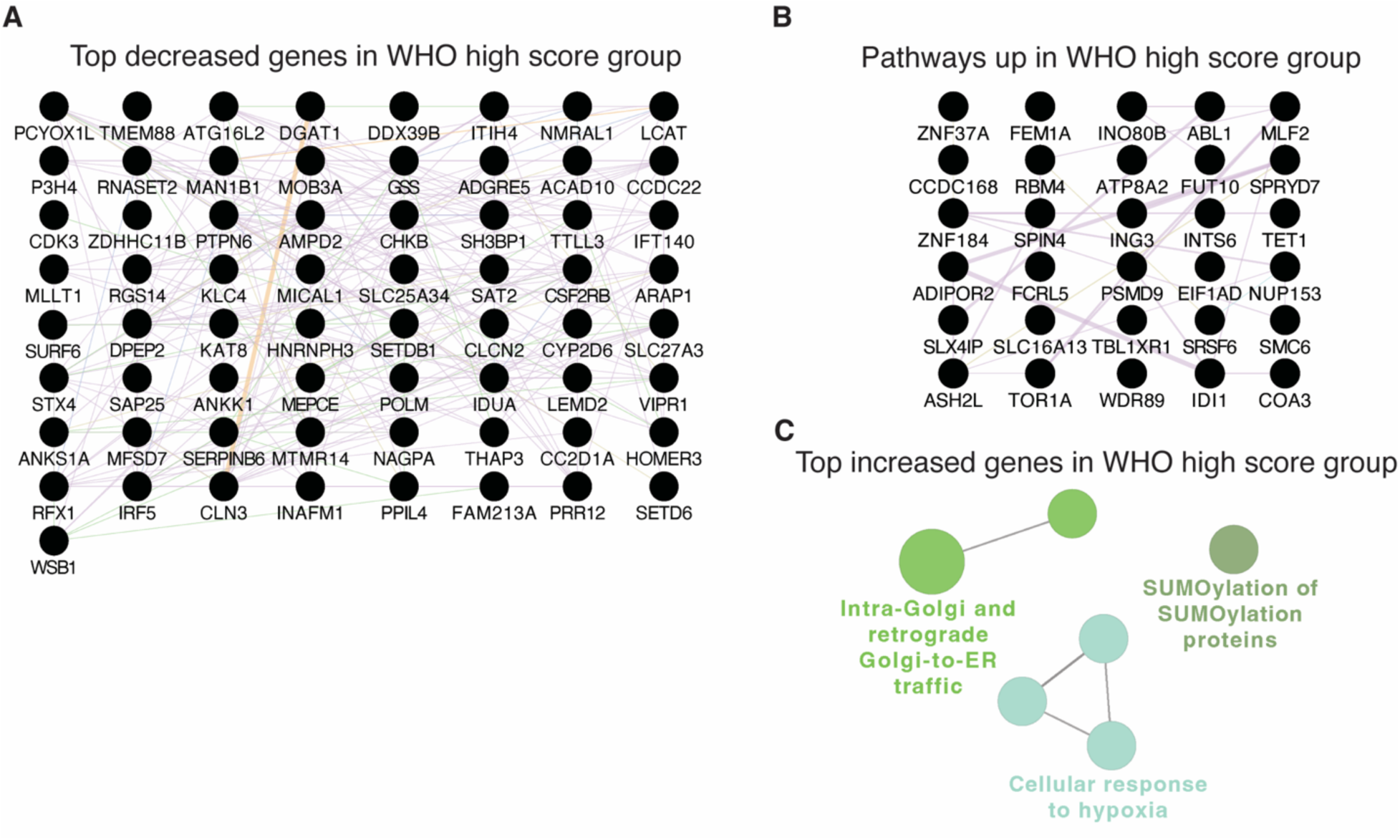
Top differentially expressed genes in COVID-19 high vs low disease severity groups. (A) 65 genes which were significantly (adjusted p<0.05) increased in participants with low disease severity (WHO score <5). (B) 30 genes which were significantly (adjusted p<0.05) increased in participants with high disease severity (WHO score >5). (C) Network-based pathways enrichment using ClueGO identifies pathways of protein trafficking, SUMOylation and response to hypoxia as enriched for genes increased in the high COVID-19 disease severity group.

**Supplemental Figure 3.**
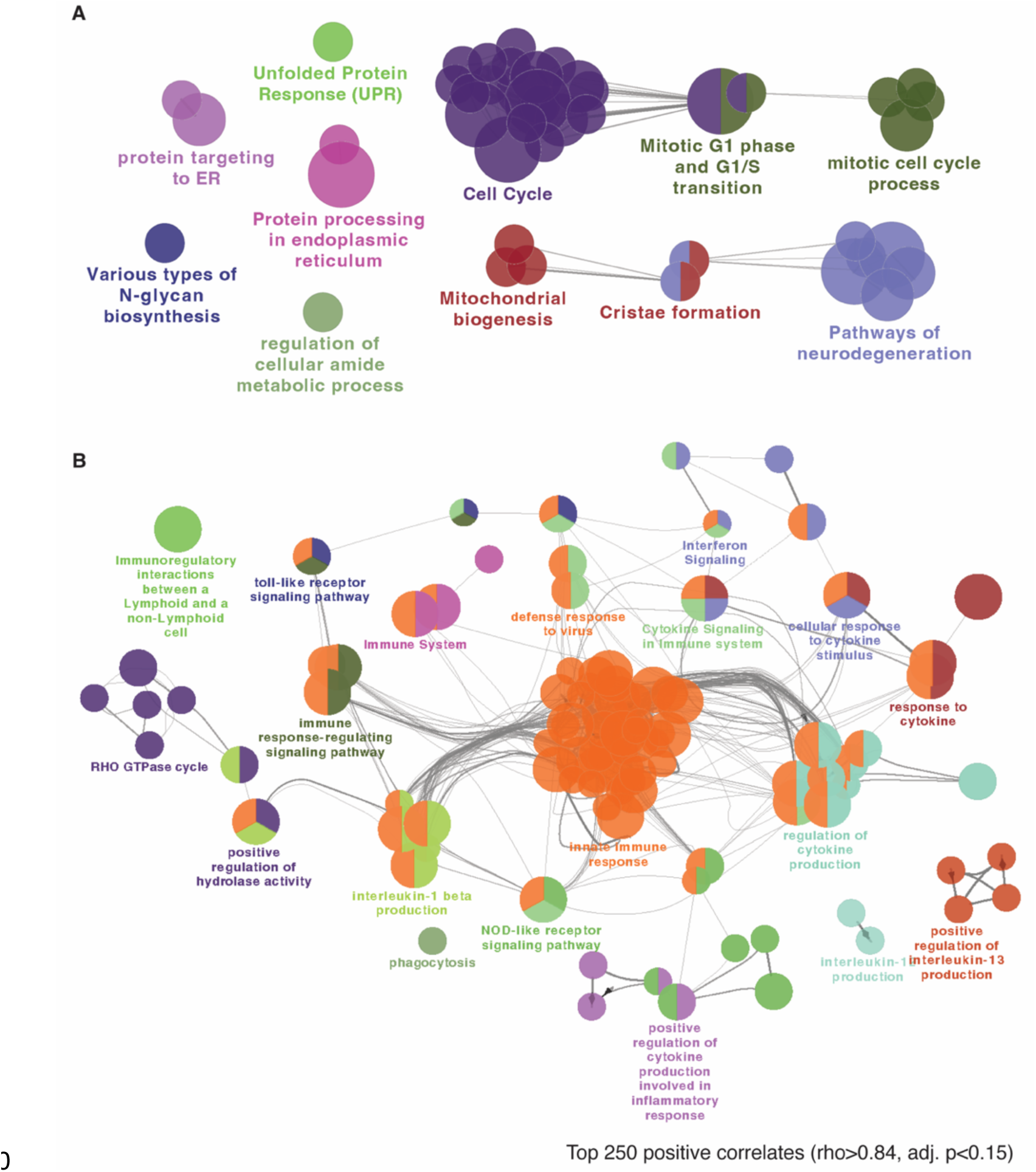
Network based pathway enrichment for the top positive and negative correlates of EPA. Network-based pathway enrichment was performed using ClueGO with (A) the top 250 positively correlating genes with EPA [rho≥0.80, adj. p<0.15] and (B) the top 250 negatively correlating genes with EPA [rho≤-0.84, adj. p<0.15]. This analysis confirmed pathways of cell cycling and division as enriched for genes positively correlated with EPA levels in plasma. Pathways of innate immunity, antiviral responses, cytokine signaling and RHO-GTPase activity were found to be enriched for genes which negatively correlated with EPA levels in plasma.

**Supplemental Figure 4:**
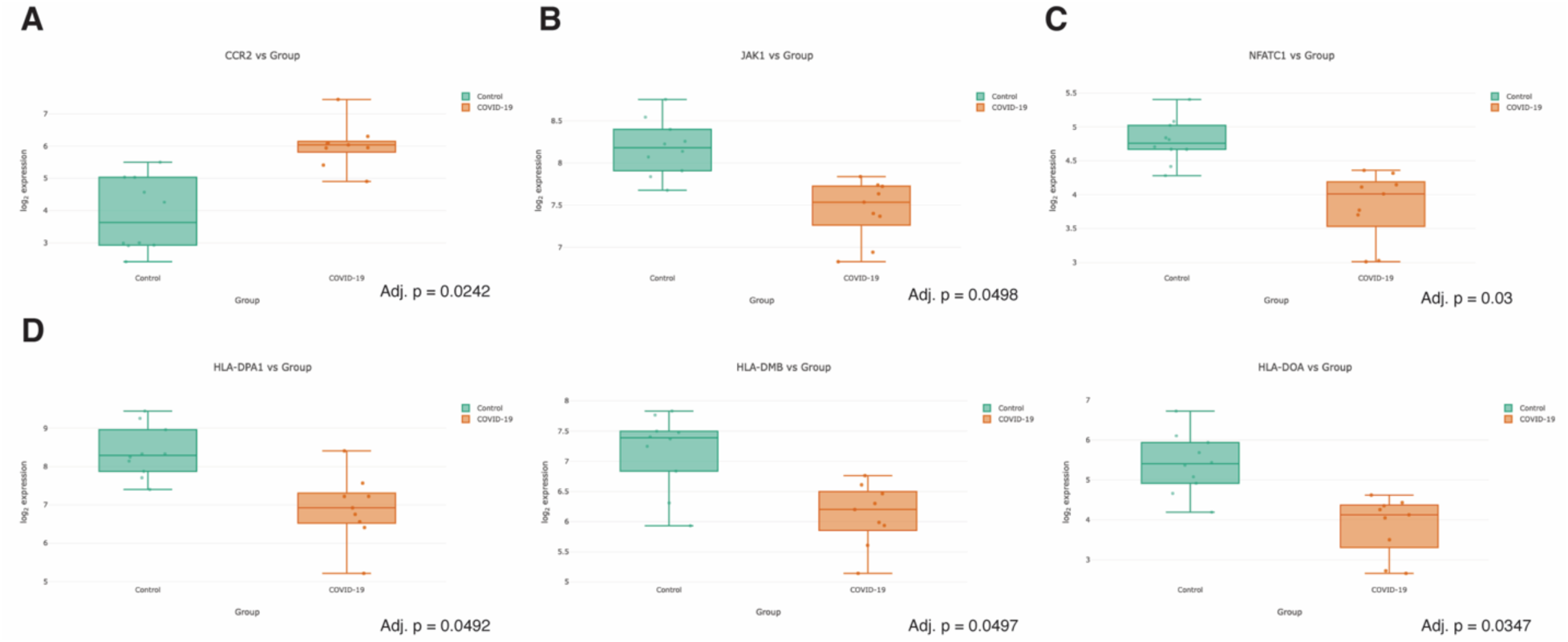
Select genes which are significantly modulated in healthy control compared to COVID-19 samples. Boxplots showing the distribution of selected significantly modulated genes between healthy control and COVID-19 samples. (A) CCR2, a chemokine receptor involved in inflammatory cell migration, is increased in COVID-19. (B) JAK1, a key kinase in STAT signaling by interferons and cytokines, (C) NFATC1, a key transcription factor in T cells, and (D) members of the MHC Class II presentation system are all significantly decreased in expression in COVID-19 samples.

**Supplemental Figure 5:**
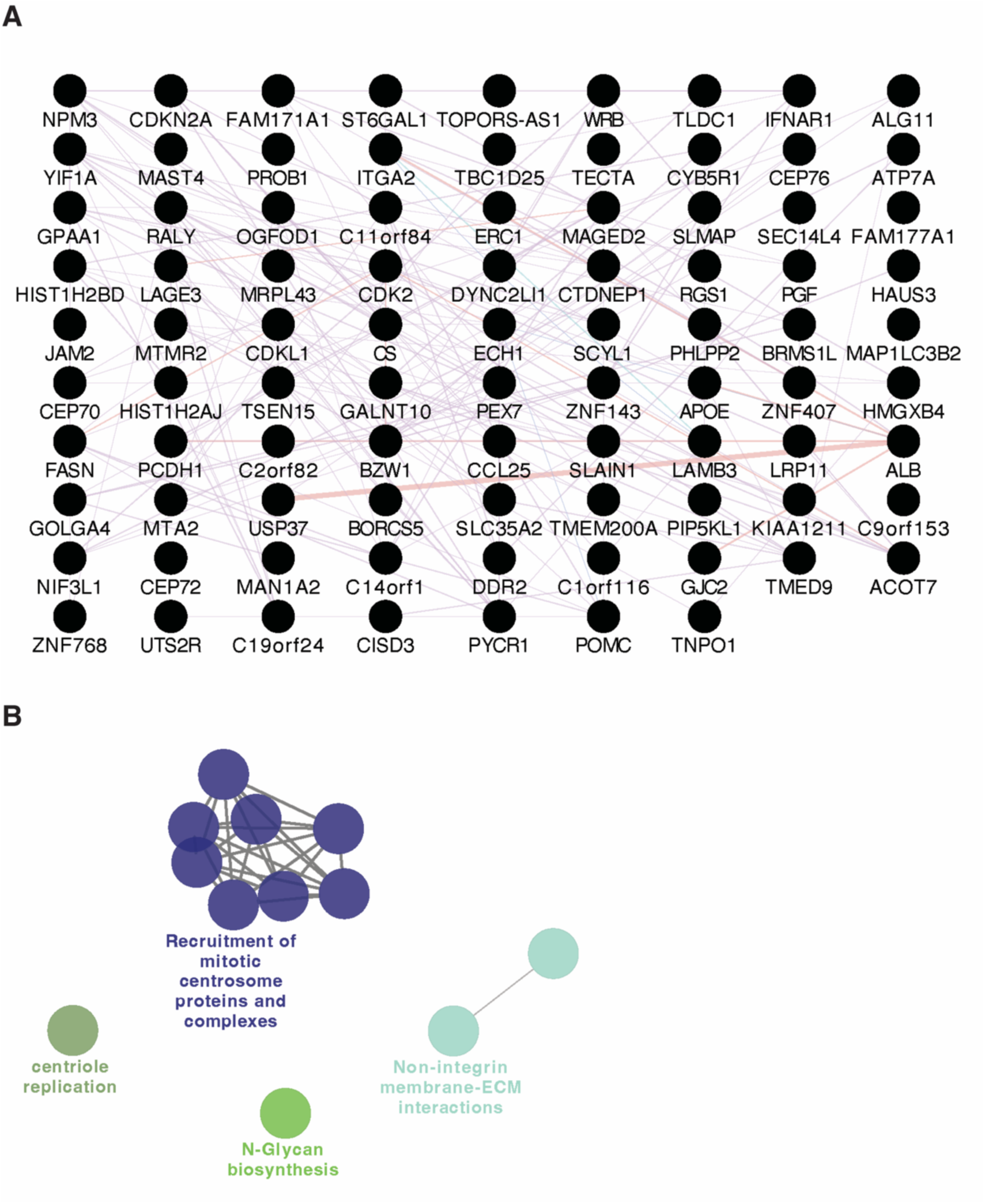
Genes which positively correlate with Eicosapentaenoic Acid (EPA). (A) Network of the 88 genes which significantly and positively correlated (p<0.05) with EPA levels. (B) Pathways showing enrichment for these significantly correlating genes using ClueGO.

## References

1. Pollard CA, Morran MP, Nestor-Kalinoski AL. The COVID-19 pandemic: a global health crisis. Physiol Genomics. 2020;52(11):549–57.

2. Dessie ZG, Zewotir T. Mortality-related risk factors of COVID-19: a systematic review and meta-analysis of 42 studies and 423,117 patients. BMC Infect Dis. 2021 Dec 1;21(1).

3. Paules CI, Marston HD, Fauci AS. Coronavirus Infections—More Than Just the Common Cold. JAMA. 2020 Feb 25;323(8):707–8.

4. Montazersaheb S, Hosseiniyan Khatibi SM, Hejazi MS, Tarhriz V, Farjami A, Ghasemian Sorbeni F, et al. COVID-19 infection: an overview on cytokine storm and related interventions. Virology Journal 2022 19:1. 2022 May 26;19(1):1–15.

5. Selva KJ, Chung AW. Insights into how SARS-CoV2 infection induces cytokine storms. Trends Immunol. 2022 Jun 1;43(6):417–9.

6. Mokhtari T, Hassani F, Ghaffari N, Ebrahimi B, Yarahmadi A, Hassanzadeh G. COVID-19 and multiorgan failure: A narrative review on potential mechanisms. J Mol Histol. 2020 Dec 1;51(6):613.

7. Lopes-Pacheco M, Silva PL, Cruz FF, Battaglini D, Robba C, Pelosi P, et al. Pathogenesis of Multiple Organ Injury in COVID-19 and Potential Therapeutic Strategies. Front Physiol. 2021 Jan 28;12:593223.

8. Dennis EA, Norris PC. Eicosanoid storm in infection and inflammation. Nat Rev Immunol. 2015 Aug 25;15(8):511–23.

9. Yoshikai Y. Roles of prostaglandins and leukotrienes in acute inflammation caused by bacterial infection. Curr Opin Infect Dis. 2001;14(3):257–63.

10. Zaid Y, Doré É, Dubuc I, Archambault AS, Flamand O, Laviolette M, et al. Chemokines and eicosanoids fuel the hyperinflammation within the lungs of patients with severe COVID-19. J Allergy Clin Immunol. 2021 Aug 1;148(2):368–380.e3.

11. Irún P, Gracia R, Piazuelo E, Pardo J, Morte E, Paño JR, et al. Serum lipid mediator profiles in COVID-19 patients and lung disease severity: a pilot study. Sci Rep. 2023 Dec 1;13(1).

12. Basil MC, Levy BD. Specialized pro-resolving mediators: endogenous regulators of infection and inflammation. Nat Rev Immunol. 2016 Jan 1;16(1):51–67.

13. Kumar V, Yasmeen N, Chaudhary AA, Alawam AS, Al-Zharani M, Suliman Basher N, et al. Specialized pro-resolving lipid mediators regulate inflammatory macrophages: A paradigm shift from antibiotics to immunotherapy for mitigating COVID-19 pandemic. Front Mol Biosci. 2023 Feb 3;10.

14. Koenis DS, Beegun I, Jouvene CC, Aguirre GA, Souza PR, Gonzalez-Nunez M, et al. Disrupted Resolution Mechanisms Favor Altered Phagocyte Responses in COVID-19. Circ Res. 2021 Aug 6;129(4):E54–71.

15. Hayashi JY, Simizo A, Miyamoto JG, Costa LVS, Souza OF, Chiarelli T, et al. Humoral and cellular responses to vaccination with homologous CoronaVac or ChAdOx1 and heterologous third dose with BNT162b2. J Infect. 2022 Jun 1;84(6):834.

16. Liang W, Gu M, Zhu L, Yan Z, Schenten D, Herrick S, et al. The main protease of SARS-CoV-2 downregulates innate immunity via a translational repression. Signal Transduct Target Ther. 2023 Dec 1;8(1).

17. Halwani R, Pulvirenti F, Al-Muhsen S. Editorial: Dysregulation of immunity predisposing to severe COVID-19 infection. Front Immunol. 2022 Dec 2;13.

18. Chu J, Xing C, Du Y, Duan T, Liu S, Zhang P, et al. Pharmacological inhibition of fatty acid synthesis blocks SARS-CoV-2 replication. Nature Metabolism 2021 3:11. 2021 Sep 27;3(11):1466–75.

19. Zhao T, Wang C, Duan B, Yang P, Wu J, Zhang Q. Altered Lipid Profile in COVID-19 Patients and Metabolic Reprogramming. Front Microbiol. 2022 May 12;13:863802.

20. Kowalska K, Sabatowska Z, Forycka J, Młynarska E, Franczyk B, Rysz J. The Influence of SARS-CoV-2 Infection on Lipid Metabolism—The Potential Use of Lipid-Lowering Agents in COVID-19 Management. Biomedicines. 2022 Sep 1;10(9).

21. Irún P, Gracia R, Piazuelo E, Pardo J, Morte E, Paño JR, et al. Serum lipid mediator profiles in COVID-19 patients and lung disease severity: a pilot study. Scientific Reports 2023 13:1. 2023 Apr 20;13(1):1–14.

22. Norris PC, Libreros S, Chiang N, Serhan CN. A cluster of immunoresolvents links coagulation to innate host defense in human blood. Sci Signal. 2017 Aug 1;10(490).

23. Regulska M, Szuster-Głuszczak M, Trojan E, Leśkiewicz M, Basta-Kaim A. The Emerging Role of the Double-Edged Impact of Arachidonic Acid-Derived Eicosanoids in the Neuroinflammatory Background of Depression. Curr Neuropharmacol. 2021 Aug 8;19(2):278–93.

24. Ziegler CGK, Miao VN, Owings AH, Navia AW, Tang Y, Bromley JD, et al. Impaired local intrinsic immunity to SARS-CoV-2 infection in severe COVID-19. Cell. 2021 Sep 2;184(18):4713–4733.e22.

25. Löfgren L, Forsberg GB, Ståhlman M. The BUME method: a new rapid and simple chloroform-free method for total lipid extraction of animal tissue. Scientific Reports 2016 6:1. 2016 Jun 10;6(1):1– 11.

26. Quehenberger O, Armando AM, Dennis EA. High sensitivity quantitative lipidomics analysis of fatty acids in biological samples by gas chromatography-mass spectrometry. Biochim Biophys Acta. 2011 Nov;1811(11):648–56.

27. Shannon P, Markiel A, Ozier O, Baliga NS, Wang JT, Ramage D, et al. Cytoscape: a software environment for integrated models of biomolecular interaction networks. Genome Res. 2003;13(11):2498–504.

28. Dobin A, Davis CA, Schlesinger F, Drenkow J, Zaleski C, Jha S, et al. STAR: ultrafast universal RNA-seq aligner. Bioinformatics. 20121025th ed. 2013;29(1):15–21.

29. Robinson MD, Oshlack A. A scaling normalization method for differential expression analysis of RNA-seq data. Genome Biol. 20100302nd ed. 2010;11(3):R25.

30. Law CW, Chen Y, Shi W, Smyth GK. voom: Precision weights unlock linear model analysis tools for RNA-seq read counts. Genome Biol. 20140203rd ed. 2014;15(2):R29.

31. Ritchie ME, Phipson B, Wu D, Hu Y, Law CW, Shi W, et al. limma powers differential expression analyses for RNA-sequencing and microarray studies. Nucleic Acids Res. 20150120th ed. 2015;43(7):e47.

32. Mostafavi S, Ray D, Warde-Farley D, Grouios C, Morris Q. GeneMANIA: a real-time multiple association network integration algorithm for predicting gene function. Genome Biol. 20080627th ed. 2008;9 Suppl 1(Suppl 1):S4.

33. Bindea G, Mlecnik B, Hackl H, Charoentong P, Tosolini M, Kirilovsky A, et al. ClueGO: a Cytoscape plug-in to decipher functionally grouped gene ontology and pathway annotation networks. Bioinformatics. 20090223rd ed. 2009;25(8):1091–3.

34. Harris MA, Clark J, Ireland A, Lomax J, Ashburner M, Foulger R, et al. The Gene Ontology (GO) database and informatics resource. Nucleic Acids Res. 2004;32(Database issue):D258–61.

35. Kanehisa M, Goto S. KEGG: kyoto encyclopedia of genes and genomes. Nucleic Acids Res. 2000;28(1):27–30.

36. Fabregat A, Jupe S, Matthews L, Sidiropoulos K, Gillespie M, Garapati P, et al. The Reactome Pathway Knowledgebase. Nucleic Acids Res. 2018;46(D1):D649–d655.

37. Halade G V, Kain V, Hossain S, Parcha V, Limdi NA, Arora P. Arachidonate 5-lipoxygenase is essential for biosynthesis of specialized pro-resolving mediators and cardiac repair in heart failure. Am J Physiol Heart Circ Physiol. 2022/08/27. 2022;323(4):H721–h737.

38. Lipman D, Safo SE, Chekouo T. Multi-omic analysis reveals enriched pathways associated with COVID-19 and COVID-19 severity. PLoS One. 2022 Apr 1;17(4).

39. Wu P, Chen D, Ding W, Wu P, Hou H, Bai Y, et al. The trans-omics landscape of COVID-19. Nat Commun. 2021 Dec 1;12(1).

40. Overmyer KA, Shishkova E, Miller IJ, Balnis J, Bernstein MN, Peters-Clarke TM, et al. Large-Scale Multi-omic Analysis of COVID-19 Severity. Cell Syst. 2021 Jan 20;12(1):23–40.e7.

41. Caterino M, Gelzo M, Sol S, Fedele R, Annunziata A, Calabrese C, et al. Dysregulation of lipid metabolism and pathological inflammation in patients with COVID-19. Scientific Reports 2021 11:1. 2021 Feb 3;11(1):1–10.

42. Palmas F, Clarke J, Colas RA, Gomez EA, Keogh A, Boylan M, et al. Dysregulated plasma lipid mediator profiles in critically ill COVID-19 patients. PLoS One. 2021/08/27. 2021;16(8):e0256226.

43. Archambault AS, Zaid Y, Rakotoarivelo V, Turcotte C, Doré É, Dubuc I, et al. High levels of eicosanoids and docosanoids in the lungs of intubated COVID-19 patients. The FASEB Journal. 2021 Jun 1;35(6):e21666.

44. Basil MC, Levy BD. Specialized pro-resolving mediators: endogenous regulators of infection and inflammation. Nat Rev Immunol. 2015/12/22. 2016;16(1):51–67.

45. Schwarz B, Sharma L, Roberts L, Peng X, Bermejo S, Leighton I, et al. Severe SARS-CoV-2 infection in humans is defined by a shift in the serum lipidome resulting in dysregulation of eicosanoid immune mediators. J Immunol. 2021 Jan 1;206(2):329.

